# Smartwatch Facilitated Remote Health Care for Patients Undergoing Transcatheter Aortic Valve Replacement Amid COVID-19 Pandemic

**DOI:** 10.1101/2021.05.22.21256870

**Authors:** Xianbao Liu, Jiaqi Fan, Yuchao Guo, Hanyi Dai, Jianguo Xu, Lihan Wang, Po Hu, Xinping Lin, Cheng Li, Dao Zhou, Huajun Li, Jian’an Wang

## Abstract

**BACKGROUND:** The novel coronavirus disease-2019 (COVID-19 Pandemic) has brought difficulties to the management of patients undergoing transcatheter aortic valve replacement (TAVR).

**OBJECTIVES:** This prospective, observational cohort study sought to evaluate the feasibility of a novel, virtual, and remote health care strategy for TAVR patients with smart wearable devices.

**METHODS:** A total of 100 consecutive severe aortic stenosis patients who underwent elective transfemoral TAVR were enrolled and each received a HUAWEI smartwatch at least one day before TAVR. Vital signs were continuously tracked and recorded. Single lead electrocardiogram (ECG) was recorded periodically after TAVR. A designated heart team member was to provide remote data-assisted health care to address the medical demand.

**RESULTS:** Thirty-eight cardiac events were reported in 34 patients after discharge, with most of the events (76.0%) were detected and confirmed by the smartwatch. Six patients were advised and readmitted to the hospital for arrhythmia events, among whom, four received pacemaker implantations. The remaining 28 (82.4%) patients received telemedicine monitoring instead of face-to-face clinical visits, and three of them received new medication treatment under a doctor’s online guidance of doctors. New-onset LBBB was found in 48 patients with transient and recovered spontaneously in 30 patients, while new-onset atrial fibrillation in 4 patients. There were no significant differences in the average weekly heart rates, the ratio of abnormal or low oxygen saturation when compared with the baseline. The average daily steps increased over time significantly (baseline, 870±1353 steps; first week, 1986±2406 steps; second week, 2707±2716 steps; third week, 3059±3036 steps; fourth week, 3678±3485 steps, p < 0.001).

**CONCLUSIONS:** Smartwatch can facilitate remote health care for patients undergoing TAVR during COVID-19 and enables a novel remote follow-up strategy. The majority of cardiac clinical events that occurred within 30-day follow-up were detected by the smartwatch, mainly due to the record of conduction abnormality. (SMART Watch Facilitated Early Discharge in Patients undergoing Transcatheter Aortic Valve Replacement, NCT04454177).

## Introduction

Coronavirus disease (COVID-19) has rapidly spread around the world in 2020 as a novel respiratory infectious disease. It threatens public health severely, causing over 140,000,000 infections and 3,000,000 deaths up to date (04-18-2021). Shorter length of stay for surgical patients facilitates the implementation of reducing medical contact as much as it can to slow down COVID-19 spread. It poses grave challenges to the traditional post-operative management, especially that of frail and old patients such as patients with severe aortic stenosis (AS).

Huawei Watch GT 2 Pro is a wearable smartwatch that could track health information including continuing 1-lead electrocardiography (ECG), blood oxygen saturation, heart rate, and steps. It can transmit data to most smartphones and upload it to the medical center in time. Transcatheter aortic valve replacement (TAVR) is a minimally invasive treatment of severe AS, which can significantly shorten the in-hospital time and is recommended to all risk profile AS patients in the 2020 ACC/AHA updated guideline (1). However, late postoperative complications, including late conduction disturbances (≥48 h), remain life-threatening complications and are impossible to be detected in all patients before discharge, especially for those who receive next-day or early-discharged patients. Moreover, the pandemic has caused more failures in detecting these late complications. Yet, mobile ECG and vital sign monitoring devices are keys to this issue. However, most prolonged ECG and vital-sign monitoring devices on market are wire connected or invasively implanted, which means they are inconvenient and risky for patients’ daily life. Therefore, the study was performed to evaluate the first clinical application of this promising device to provide remote health care for patients after the TAVR procedure.

## Methods

### Study design and patient population

The SMART TAVR (SMART Watch Facilitated Early Discharge in Patients undergoing Transcatheter Aortic Valve Replacement) registry (NCT04454177) is a prospective cohort study in China. The study protocol was approved by the medical ethics committee of the Second Affiliated Hospital of Zhejiang University and was in accordance with the Declaration of Helsinki. All participants provided informed consent to participating in the study before the procedure.

Participants received a smartwatch paired to their smartphone within 24 hours before the procedure. A smartwatch was employed to record the following parameters: heart rate, steps, pulse oxygen saturation, and single lead electrocardiograph (ECG) after triggering. They were instructed how to perform ECGs using the smartwatch and transmit smartwatch sensor data via the smartphone application to a web-based clinical research platform. ECG measurements were required to be at least 15 seconds long to ensure quality and accuracy.

The registry was conducted between July 2020 and March 2021 during the pandemic and a total of 100 consecutive patients undergoing elective transfemoral TAVR were enrolled. The purpose was to reduce the face-to-face medical contact through smartwatches during follow-up for post-TAVR patients amid COVID-19 pandemic. The major exclusion criteria were: 1) Severe complications of TAVR, such as death, and conversion to SAVR; 2) Life expectancy is less than 12 months due to non-heart disease (such as cancer, chronic liver disease, chronic kidney disease, or chronic end-stage lung disease, etc.); 3) Severe dementia (cannot sign research informed consent, cannot take care of themselves or cooperate in the study visit); 4) The investigator believes that the patient is not suitable to participate in the study or complete the follow-up prescribed by the protocol from other medical, social and psychological aspects; 5) The patient is currently participating in another randomized study. All patients finished a 30-day follow-up.

Patients primarily diagnosed by the transthoracic echocardiogram (TTE) as severe aortic stenosis underwent multi-slice computed tomography angiography (MDCT). MDCT was crucial to assess anatomical features of the patients and was played a vital role in formulating TAVR strategy. The decision on device and operation approach was made based on 3mensio software (3mensio Medical Imaging BV, Bilthoven, the Netherlands) as previously described. The minimal luminal diameter feasible for femoral artery access was 6mm theoretically. Risk assessment of our study was assisted by the Society of Thoracic Surgeons (STS) score gained preoperatively. Dominant Valves implanted in the procedure were Venus A Series (Venus A and Venus A Plus, Venus Medtech, Hangzhou, China). VitaFlow Series (VitaFlow I and VitaFlow II, Microport, Shanghai, China) and SAPIEN 3 valves (Edwards Lifesciences, Irvine, California) were also applied. Details of TAVR procedures have been previously reported. All TAVR procedures were performed under local anesthesia with sedation, which contributed to early mobilization and early discharge. Temporary pacemakers were implanted in all patients during the procedure. ProGlides were used for the puncture site closure to reduce the access site-related complications to facilitate early discharge.

Patients were transferred to the cardiac intensive care unit after the operation and received intensive monitoring. All femoral lines were removed at the end of operation except temporary pacemaker and racial artery catheterization was removed 4 hours after -TAVR. The temporary pacemaker would be removed off within 24 hours in the absence of new-onset or aggravating conduction disturbance. Permanent pacemaker implantation was considered in the case of a high-grade atrioventricular block or long intervals occurred.

### Data collection

Data collection included baseline characteristics, procedural data, predischarge outcomes, and follow-up data. Baseline clinical characteristics medical comorbidities and medicines patients were taking were collected the day before the procedure. Peri-procedural complications were defined according to Valve Academic Research Consortium-2 criteria. During hospitalization, ECGs were collected within 24 hours before the procedure, immediately following the procedure, 4 hours following the procedure, and subsequently every day during the hospitalization to determine whether there was a conduction disturbance. Follow-up data consist of patients-reported clinical events, smartwatch sensor data, and examination at 30-day follow-up. Patients were required to submit smartwatch biometric data, particularly the ECG data, twice per day in the week following TAVR discharge and at least two days a week for the subsequent month after TAVR discharge. If compliance declined, patients would be reminded by a member of the cardiac research team. Furthermore, patients could report their symptoms, such as chest pain, palpitation, and syncope, recorded and record and transmit single-lead ECGs through the smartphone app simultaneously. Experienced electrophysiologists in the ECG Core-Lab of this study interpreted ECG on a daily basis to avoid wrong diagnosis and treatment of patients. Patients would be required to seek emergent treatment if necessary. The clinical event committee reviewed and discussed all clinical events regularly to confirm the outcomes in the presented study. The data and safety monitoring board would review data from the ongoing trial and ensured that participants were not exposed to undue risk. Patients were followed up by a face-to-face assessment 30 days after the procedure.

### Statistical analysis

All statistical analyses were computed using SPSS 25 (IBM SPSS, USA). Kolmogorov-Smirnov test was used to analyze the distribution of continuous variables. Continuous variables expressed as mean ± SD and were compared with Student’s t-test or Mann-Whitney U test based on their distribution. Categorical data were presented as frequency (percentage) and compared with the chi-square test or Fisher exact test. A two-side p < 0.05 was considered statistically significant.

## Results

### Baseline characteristics

Amid COVID-19 pandemic period, a total of 100 consecutive patients who underwent TAVR between July 16, 2020, and Mar 16, 2021 received remote health care assisted with Huawei GT Pro 2 watches and were included in the study. No patients withdrew from the study and all patients finished 30-day follow-up. The average age of the enrolled patients was 73.1 ± 7.6 and 55.0% of them were men. There were overall 71% of the patients had NYHA class III or IV symptoms before the TAVR procedure. Detailed baseline characteristics were provided in Table 1.

**Table 1.** Baseline characteristics.

|  | **Number of patients, n = 100** |
| --- | --- |
| **Age, yrs** | 73.1±7.6 |
| **Male, n(%)** | 55(55.0) |
| **BMI, kg/m<sup>2</sup>** | 23.42±3.74 |
| **NYHA III/IV, n(%)** | 71(71.0) |
| **STS score, %** | 4.35±3.25 |
| **Prior pacemaker, n(%)** | 3(3.0) |
| **Hypertension, n(%)** | 56(56.0) |
| **Diabetes mellitus, n(%)** | 21(21.0) |
| **COPD, n(%)** | 27(27.0) |
| **Prior PCI, n(%)** | 12(12.0) |
| **Prior electrocardiography** |  |
| RBBB, n(%) | 4(5.0) |
| LBBB, n(%) | 2(2.5) |
| First-degree AV block, n(%) | 10(12.5) |
| Atrial fibrillation/atrial flutter, n(%) | 14(17.5) |
| CHADS-VASc Score | 2.9±1.2 |
| HAS-BLED | 1.8±1.0 |
| Anticoagulation, n(%) | 6(7.5) |
| **Echocardiography** |  |
| AVA, cm<sup>2</sup> | 0.67±0.25 |
| Mean Gradient, mmHg | 55.4±22.2 |
| Maximum velocity, m/s | 4.72±1.11 |
| LVEF, % | 59.0±10.5 |

Data are presented as mean ± SD or no. (%).
AVA, aortic valve area; BMI, body mass index; COPD, chronic obstructive pulmonary disease; LBBB, left bundle branch block; LVEF, left ventricular ejection fraction; RBBB, right bundle branch block; STS, Society of Thoracic Surgeons.

### Procedural characteristics and clinical outcome

All patients underwent transfemoral TAVR procedure among whom 91% were implanted with a self-expanding valve and 9% patients underwent balloon expanding TAVR. Four patients underwent second valve implantation and no patients were transferred to surgical valve replacement. Eighty-two percent of patients were discharged early, among whom, 56% were discharged the next day after the surgery. No death or myocardial infarction occurred during follow-up. The rate of stroke was 1.0% both in hospital and at 30-day follow-up. All patients had NYHA class I or II functions at 30-day follow-up. The transvalvular mean gradient was 11.7 ± 5.3 mmHg and the mean prosthetic valve area was 1.63 ± 0.37 cm^2^ according to 30-day echocardiography. Procedural characteristics and clinical outcomes were presented in Table 2 and Supplementary Table 1.

**Table 2.** Clinical outcome in-hospital and during 30-day follow-up.

|  | Number of patients, n = 100 |
| --- | --- |
| **In hospital** |  |
| Mortality, n(%) | 0(0.0) |
| MI, n(%) | 0(0.0) |
| Stroke, n(%) | 1(1.0) |
| Pacemaker implantation, n(%) | 10(10.0) |
| Next-day discharge, n(%) | 56(56.0) |
| Early-day discharge, n(%) | 82(82.0) |
| Echocardiography |  |
| AVA, cm<sup>2</sup> | 1.71±0.48 |
| Mean Gradient, mmHg | 12.4±5.4 |
| Maximum velocity, m/s | 2.41±0.51 |
| LVEF, % | 60.2±9.1 |
| Moderate or severe paravalvular leakage, n(%) | 5(5.0) |
| **30-day follow-up** |  |
| Mortality, n(%) | 0(0.0) |
| MI, n(%) | 0(0.0) |
| Stroke, n(%) | 1(1.0) |
| Pacemaker implantation, n(%) | 4(4.0) |
| Rehospitalization, n(%) | 11(11.0) |
| Cardiac Rehospitalization, n(%) | 8(8.0) |
| Cardiac rehospitalization by watch, n(%) | 6(6.0) |
| Echocardiography |  |
| AVA, cm<sup>2</sup> | 1.63±0.37 |
| Mean Gradient, mmHg | 11.7±5.3 |
| Maximum velocity, m/s | 2.32±0.56 |
| LVEF, % | 59.9±9.5 |
| Moderate or severe paravalvular leakage, n(%) | 5(5.0) |

Data are presented as mean ± SD or no. (%).
AVA, aortic valve area; LVEF, left ventricular ejection fraction; MI, myocardial infarction.

### The clinical event, arrhythmia findings, and biometric parameters detected by the smartwatch

The clinical event, arrhythmia findings, and biometric parameters recorded by the watch during 30-day follow-up were provided in Table 3. Thirty-eight cardiac clinical events were reported in 34 patients after discharge. Most of these events (76.0%) were detected and confirmed by the smartwatch, leading to six patients readmitted to the hospital. The remaining 28 patients received telemedicine monitoring, and three of them received new medication treatment under a doctor’s online guidance of doctors. Therapeutic changes occurred in nine patients, among whom, four received pacemaker implantation and five medical therapy changes (β blocker in three patients, potassium treatment in one, and amiodarone in one). The therapeutic changes in nine patients were presented in Supplementary Table 2.

**Table 3.**
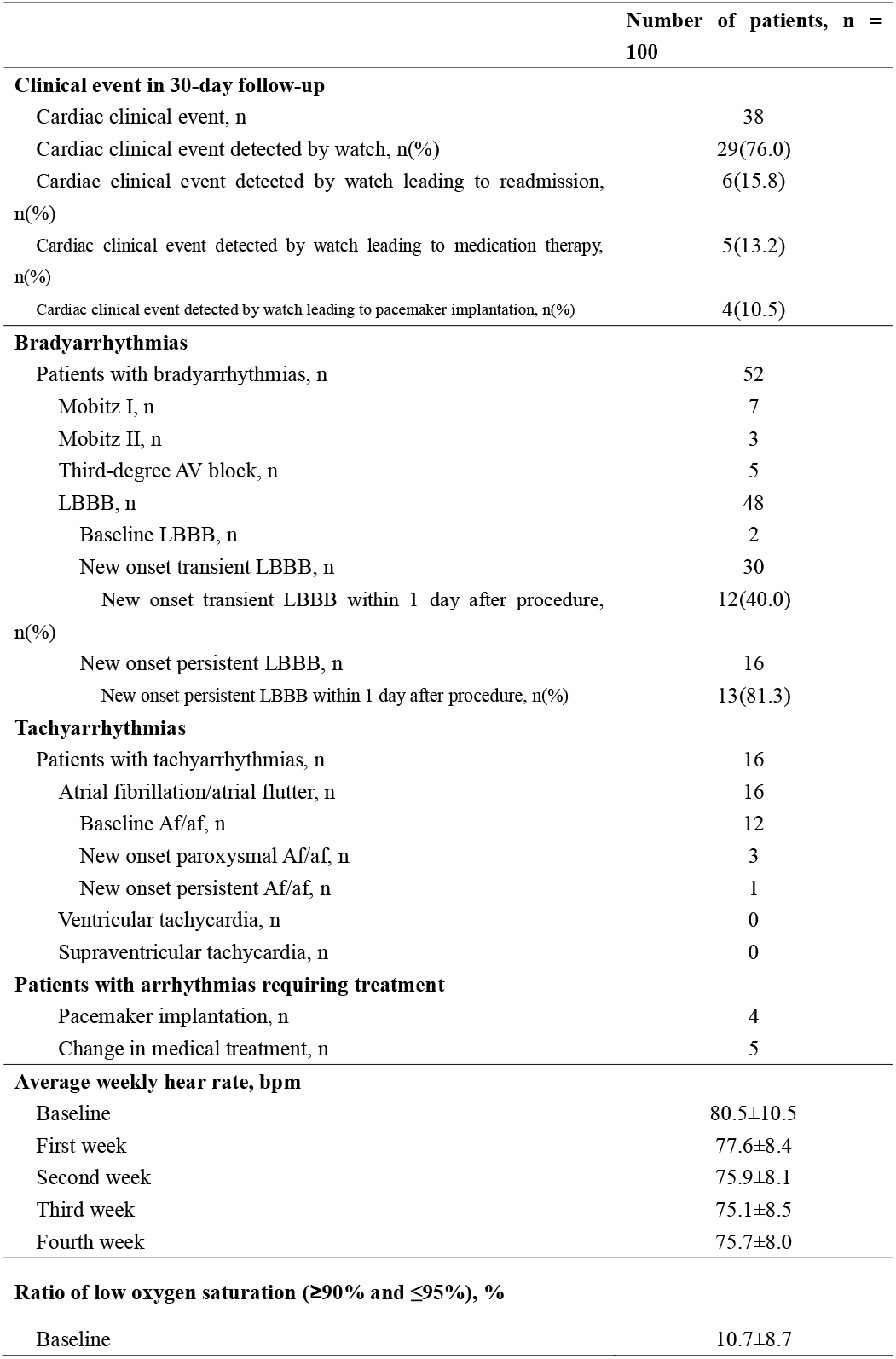
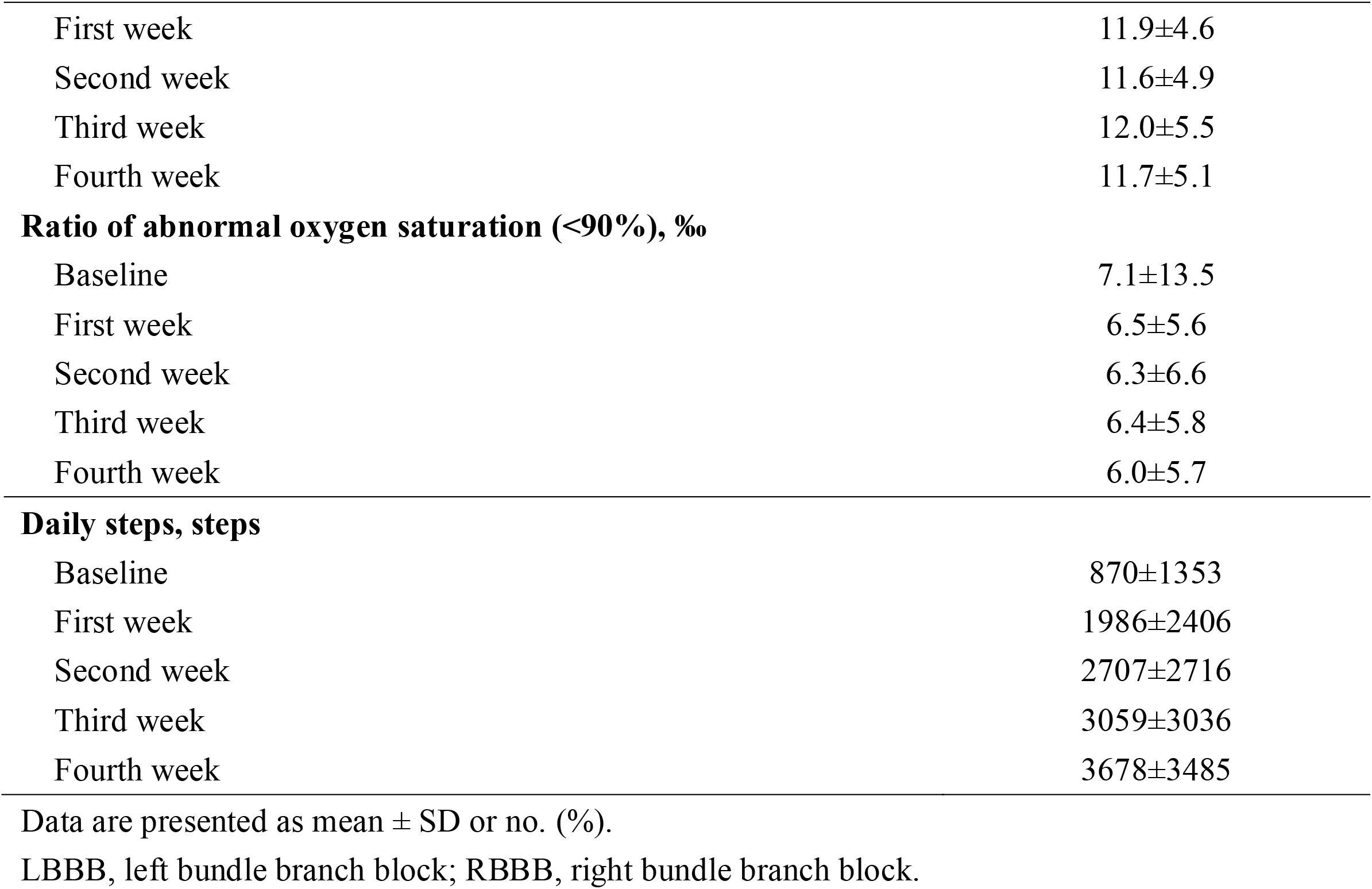
Clinical event and biometric parameters recorded by watch during 30-day follow-up.

The incidence of bradycardia and tachycardiac arrhythmia was diagnosed in 52% and 16% of patients respectively from baseline to 30-day follow-up. Among 52 patients with bradycardic arrhythmia detected by the smartwatch, LBBB was found in 48 patients, five patients suffered complete heart block, and three developed second-degree atrioventricular block type 2 (Mobitz II). LBBB showed transient characteristics and recovered spontaneously in 30 patients, while persistent LBBB was found in 16 patients. Most of the new-onset persistent LBBB (81.3%) occurred on the first day after the procedure while only 40.0% transient LBBB could be found in this period. As for tachycardiac events, atrial fibrillation (AF) or atrial flutter (AFL) were found in 16 patients, the majority of whom (75.0%) had preoperative atrial fibrillation or flutter (Table 3). No other tachycardiac events such as severe ventricular or supraventricular tachycardia were found by ECG in the hospital or by the smartwatch. In total, Permanent pacemakers were implanted in 14 patients, 10 of whom received pacemaker implantation before discharge and four received pacemaker implantations during 30-day follow-up.

The heart rates, blood oxygen, and steps were recorded by the smartwatch on a real-time basis every day and were uploaded regularly through smartphone. The data were available to all patients. The daily steps counted by smartwatch were found to increase over the period time from baseline to 30-day follow-up (p < 0.001). The mean reported steps in four weeks were 1986±2406, 2707±2716, 3059±3036, 3678±3485, respectively, while 870±1353 steps at baseline. The average weekly heart rates in the first four weeks were 77.6±8.4, 75.9±8.1, 75.1±8.5, 75.7±8.0, though no trend was found over time. The ratio of abnormality or low oxygen saturation in four weeks were 6.5±5.6‰, 6.3±6.6‰, 6.4±5.8‰, 6.0±5.7‰ and 11.9±4.6‰, 11.6±4.9%, 12.0±5.5%, 11.7±5.1% respectively. There was also no trend found in oxygen saturation during follow-up. The biometric parameters recorded by the smartwatch were presented in Table 3 and Figure 1.

**Figure 1.**
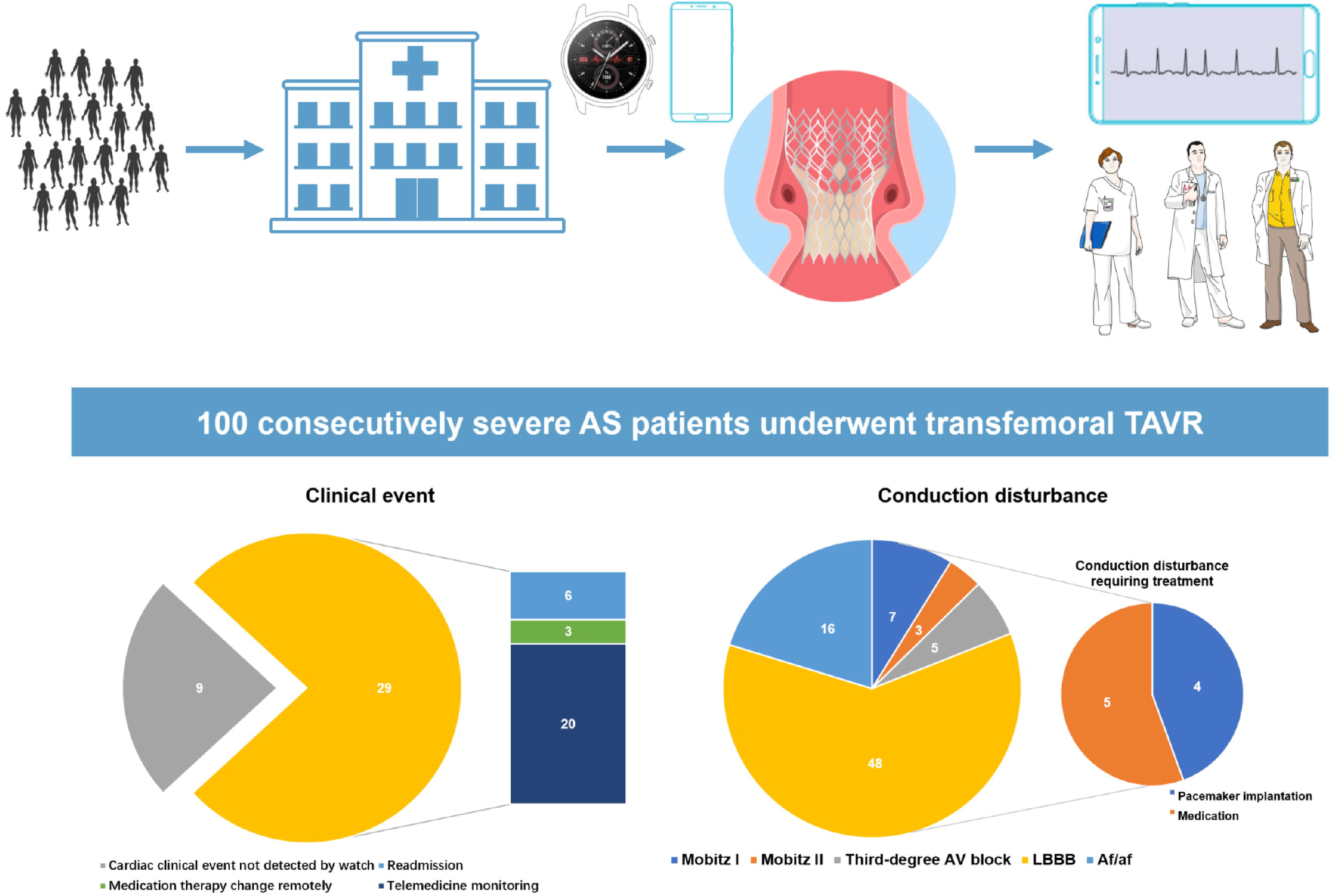
biometric parameters recorded by smartwatch at baseline and after discharge. Central illustration. (Top) Patterns of smartwatches facilitated remote health care during COVID-19 for patients undergoing TAVR. (Bottom left) Clinical event during 30-day follow-up with 29 events found by the smartwatch, 6 patients rehospitalization, 3 patients received new medication remotely, and the remaining 20 patients were in close telemedicine monitoring. (Bottom right) Conduction disturbance was detected by smartwatch after TAVR, with 48 LBBB patients, 16 atrial fibrillation patients, 7 Mobitz I patients, 3 Mobitz II patients, and 5 third-degree atrioventricular block patients. Of these patients, 5 patients received medication therapy and 4 patients received pacemaker implantation after discharge with 30-day follow-up.

## Discussion

The main findings of our study are the following: 1) The Huawei smartwatch facilitated post-discharge monitoring can reduce post-procedural medical contact during the COVID-19 epidemic. 2) Smartwatch facilitated early discharge plan is feasible without a significant increase in mortality, clinical outcome, or readmission rates. 3) Post-TAVR telehealth and monitoring with Huawei smartwatch provide a promising new way to promote remote outpatient management.

### Need of remote health care during COVID-19 pandemic

Next-day or early discharge after TAVR has been validated in multiple studies (2)(3)(4). However, delayed atrioventricular block and other TAVR-related complications or concomitants require special attention after discharge (5). Last year, the COVID-19 pandemic impeded medical management of TAVR patients, especially the elderly patients with multiple comorbidities who are at highest risk for COVID-19 infection or poor outcome (6)(7). Those unsatisfied needs resulted in the rapid development of remote patient management with new technologies and platforms (8)(9). Herein, we introduce a novel strategy to provide remote health care for outpatients after the TAVR procedure during the COVID-19 era.

To date, ambulatory ECG monitoring devices have been implemented on TAVR patients, which have brought new insight into arrhythmia detection and control (10). Traditional Holter, external spot single-lead ECG check (event recorder, smartphone, or smartwatch), mobile cardiovascular telemetry monitoring (lead, patch, or garment based), and implantable cardiac monitor are common ambulatory ECG monitoring devices (10). Certain monitoring devices may have some disadvantages like signal quality issues, noncontinuous recording, problems of patient acceptance problems, manual trigger, and financial cost (10). Thus, a new method in the follow-up with remote health care after TAVR is direly needed.

### Clinical event detected by watch following TAVR

In the study presented in this paper, 29(76%) cardiac clinical events were detected by the watch, which led to 6 readmission, 5 medication therapy, and 4 pacemaker implantations. For patients enrolled in the SMART TAVR trial, electrocardiography (ECG) monitoring, blood oxygen saturation, heart rate monitoring, and telemedicine with smartphone app were all able to detect cardiac clinical events. A remote telemonitoring system was introduced by Mathilde C Hermans in 2018 (11). Such a system has not been tested in a real clinical entity yet. Our smartwatch facilitated post-TAVR management plan has been proved to be feasible and safe with zero mortality. With the extra support of vital-sign monitoring and remote consulting, we managed to avoid the face-to-face medical visit while distinguishing four patients who developed HAVB and were in need of pacemaker implantation. It should be noticed that the mean age of our study population was 73.1 years old and all patients showed excellent adherence, which can undoubtedly serve as a reassurance in terms of patient acceptance. During the study period, we transferred our traditional TAVR management plan to a next-day discharge program. Finally, 82% of patients were discharged early from the hospital and 56% of patients had the next day discharge. We believe this remote health care system can boost patient confidence and ensure early recognition of a potential pathophysiological change in early discharge patients, which enables remote assessment and prompts readmission if necessary.

### Arrhythmias following TAVR

As proved by the present study, Huawei Watch GT 2 Pro was able to screen out more than one-half of patients with arrhythmias after TAVR. The rate of high AVB (Mobitz II and third-degree AV block) during 30-day follow-up was 6%, and the need for pacemaker implantation for HAVB was 2%. The results of this study are consistent with previous works. MARE study found 8% of patients after TAVR experienced High AVB through the invasive implantable cardiac monitor (Reveal XT) (12). Two main studies using mobile cardiovascular telemetry monitoring (MCT) after TAVR detected 9% of patients with episodes of High AVB (13)(14). Of note, the majority (75%) of bradyarrhythmia patients were asymptomatic, which was in accordance with our finding that only 2 out of 6 High AVB patients needed the pacemaker.

On the other hand, we only found 4% of new-onset atrial fibrillation/atrial flutter during 30-day follow-up period. No ventricular or supraventricular tachycardia was recorded in this study. These results match those observed in earlier studies. The detection rate of atrial fibrillation reported by the MARE study was 9% in 30 day follow-up (12). The detection rate seems to be numerically lower in studies using mobile cardiovascular telemetry monitoring, 0% and 6% respectively (15)(16). Anticoagulation treatment was initiated in only one persistent Af/af patient which was in line with suggestions from the 2021 JACC State-of-the-Art Review (10). However, the minimum AF duration prior to start anticoagulation therapy in asymptomatic AF patients remains uncertain, especially in TAVR patients who often score high in both high CHA2DS2-VASc score (thromboembolic risk) and HAS-BLED score (major bleeding risk). Thus, the decision of when to transfer a patient to anticoagulation should be individualized.

### Future perspective

This study, although preliminary, suggests that smartwatch facilitated remote health care was feasible in real-world clinical circumstances. However, further studies are necessary to secure the clinical benefit of such telemonitoring devices.

### Study limitations

This was a single-center study, and the valves used in this study were mainly first-generation ones, which may limit generalizability. Secondly, the current version of the Huawei Watch GT Pro has to be manually initiated to start recording a single lead ECG. However, with proper pre-discharge education, we found it could be handled properly by the elderly with good tolerance. Immediate alarm generation base on real-time blood oxygen saturation and heart rate monitoring will be upgraded in the next-generation software. Third, the follow-up duration of this study was

30 days, longer-term monitoring of our cohort may provide a better understanding of telemonitoring after TAVR.

## Conclusion

Smartwatch can facilitate remote health care for patients undergoing TAVR during COVID-19 and enables a novel remote follow-up strategy. The cardiac clinical event that occurred within 30-day follow-up was almost all detected by the smartwatch, mainly due to the record of conduction abnormality.

## Data Availability

Data can only be accessed after reasonable request.

## Abbreviations

AS: aortic stenosis
HR: heart rate
LBBB: left bundle branch block
RBBB: right bundle branch block
TAVR: transcatheter aortic valve replacement
TTE: transthoracic echocardiography

## Acknowledgments

The authors sincerely thank the Huawei Heart Health Research Team for the development and optimization of the ECG algorithm, headed by Mr. Xiaoxiang He. Team members include Jiabing Yan, Wenjuan Chen, Lian Wu, and Jianhua Guo. We are grateful to Hasan Jilaihawi, from the Department of Cardiology and Cardiothoracic Surgery, NYU Langone Medical Center, New York, NY, USA, for his contribution to the scientific design of the SMART TAVR trial.

## PERSPECTIVES

### WHAT IS KNOWN?

COVID-19 curtails both in-hospital time and fewer face-to-face medical visits. It poses grave challenges to the traditional post-operative management, especially of the frail and old patients with many cardiovascular comorbidities such as severe AS patients who underwent TAVR.

### WHAT IS NEW?

The noninvasive wearable smartwatch could provide remote health information including continuing 1-lead electrocardiography (ECG), blood oxygen saturation, heart rate, and steps. This presented study was the first to use this promising device to help provide remote health care for patients after the TAVR procedure during the COVID-19 pandemic.

### WHAT IS NEXT?

Long-term monitoring should be provided and more patients should be included to better understand the value of telemonitoring in TAVR patients. Next-generation of smartwatches should be smarter and more intelligent. More patients could benefit from this novel remote health care strategy in follow-up after cardiac surgery.

---

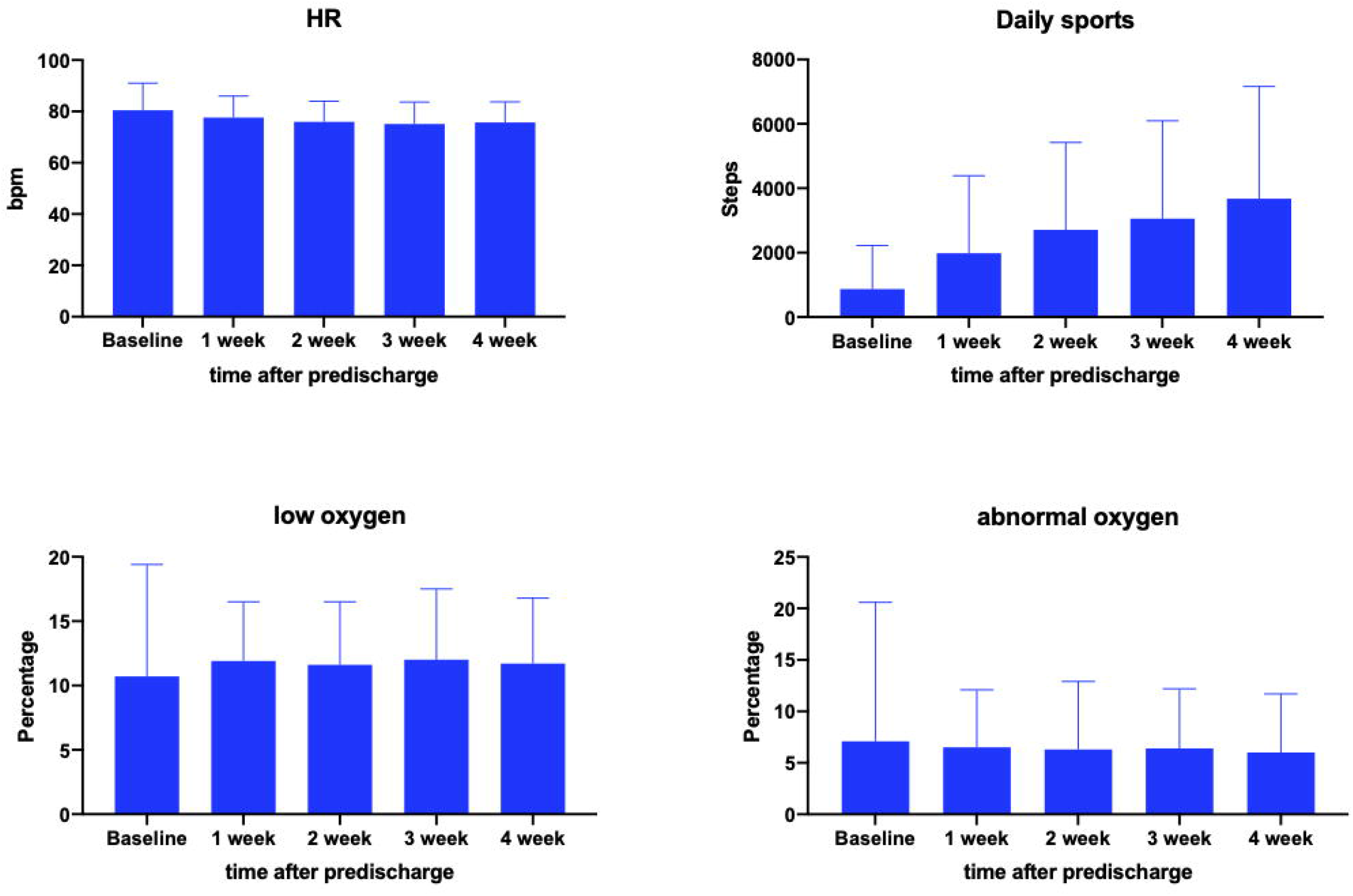

