## Supplementary Table 1 for "Smartwatch Facilitated Remote Health Care for Patients Undergoing Transcatheter Aortic Valve Replacement Amid COVID-19 Pandemic"

**Supplementary Table 1. Procedural data**

| | Number of patients, n = 100 |
|---|---|
| Second Valve, n(%) | 4(4.0) |
| Pre-dilatation, n(%) | 96(96.0) |
| THV type, n(%) | |
| Self-expanding valve, n(%) | 91(91.0) |
| Balloon expandable valve, n(%) | 9(9.0) |
| Device | |
| Early-generation device, n(%) | 65(65.0) |
| New-generation device, n(%) | 35(35.0) |
| Post-dilatation, n(%) | 63(63.0) |
| Death, n(%) | 0(0.0) |
| Myocardial infarction, n(%) | 0(0.0) |
| Aortic dissection, n(%) | 2(2.0) |
| Conversion to SAVR, n(%) | 0(0.0) |
| Annular rupture, n(%) | 0(0.0) |
| Coronary obstruction, n(%) | 0(0.0) |

Data are presented as no. (%).

SAVR, surgical aortic valve replacement; THV, transcatheter heart valve
