## Supplementary Table 2 for "Smartwatch Facilitated Remote Health Care for Patients Undergoing Transcatheter Aortic Valve Replacement Amid COVID-19 Pandemic"

### Supplementary Table 2. Conduction disturbance detected by smartwatch leading to therapy

| Patient # | Cardiovascular symptoms | Conduction Abnormalities | Detected by watch | Readmission | Therapy |
|---|---|---|---|---|---|
| 1# | cardiogenic syncope | af, LBBB, sinus arrest for 8.46 seconds | Yes | Yes | pacemaker |
| 2# | palpitations | frequent ventricular premature | Yes | No | beta-block |
| 3# | no symptoms | third-degree atrioventricular block, sinus arrest for 4.6 seconds | Yes | Yes | pacemaker |
| 4# | palpitations | frequent ventricular premature | Yes | No | potassium chloride for hypokalemia |
| 5# | dizziness, nausea | frequent ventricular premature | Yes | Yes | beta-block |
| 6# | no symptoms | high-degree atrioventricular block | Yes | Yes | pacemaker |
| 7# | dizziness | af, LBBB, prolonged RR interval | Yes | Yes | pacemaker |
| 8# | dizziness | af with hypotension | Yes | Yes | amiodarone |
| 9# | palpitations | frequent atrial premature with IVCD | Yes | No | beta-block |

af, atrial fibrillation; IVCD, intraventricular conduction disturbance
